# “Half the truth is often a great lie:” testing for tick-borne diseases and implications for surveillance

**DOI:** 10.1101/2022.02.08.22270683

**Authors:** Amanda Brown Marusiak, Brandon D. Hollingsworth, Haley Abernathy, Aidin Alejo, Victor Arahirwa, Odai Mansour, Dana Giandomenico, John Schmitz, Carl Williams, Alexis Barbarin, Ross M. Boyce

## Abstract

**Importance:** Tick-borne diseases (TBD) including Spotted Fever Group Rickettsiosis (SFGR), ehrlichiosis, and increasingly Lyme disease represent a substantial public health concern throughout much of the Southeastern United States. Yet, there is uncertainty about the epidemiology of these diseases due to pitfalls in existing diagnostic test methodologies.

**Objective:** To examine patterns of diagnostic testing and incidence of TBD in a large, academic healthcare system.

**Design:** Cross-sectional study of diagnostic test results from UNC Health for the period January 1^st^, 2017 to November 30^th^, 2020.

**Setting:** Large, academic healthcare system in central North Carolina including inpatient and outpatient facilities.

**Participants:** All Individuals seeking routine care at UNC Health facilities who had testing for SFGR, ehrlichiosis or Lyme disease performed during the study period

**Measurements:** Rates of test positivity, testing completeness, and incidence of TBD

**Results:** Among the 20,528 diagnostic tests performed, we identified 47 laboratory-confirmed, incident cases of SFGR, 27 of ehrlichiosis, and 76 of Lyme, representing incidence rates of 4.7%, 7.1%, and 0.7% respectively. However, 79.3% of SFGR tests and 74.3% *Ehrlichia* tests lacked a paired convalescent sample. The total number of tests for Lyme disease was more than SFGR and ehrlichiosis combined, despite the relatively low incidence of disease in region. Most striking, testing for ehrlichiosis was performed in only half of patients in whom SFGR was ordered, suggesting that this disease remains underrecognized. Overall, we estimate that there were 187 incident cases of SFGR and 309 of ehrlichiosis that were not identified due to incomplete testing; a number that would drastically increase – and in the case of ehrlichiosis, nearly double – the total number of cases reported.

**Conclusions and Relevance:** A majority of patients suspected of having TBD did not have testing performed in accordance with established guidelines, substantially limiting our understanding of TBD epidemiology. Furthermore, there appears to be a large discrepancy between the local burden of disease and the testing that is performed. These findings underscore the need to pursue more robust, active surveillance strategies to estimate the burden of TBDs and distribution of causative pathogens.

## Background

From 2004 to 2016, the Centers for Disease Control and Prevention (CDC) received nearly 500,000 reports of tick-borne diseases (TBD), comprising more than three-quarters of all vector-borne disease reports in the continental United States (1). Notably, reports of TBD doubled over this same time period, with consistent annual increases in Lyme disease, spotted fever group rickettsiosis (SFGR) and ehrlichiosis. These estimates, based on reports from state health departments, likely capture only a fraction of total infections and clinical illness (2-4). These trends will likely be exacerbated by global climate change, which may affect not only the geographic distribution of ticks, but also the tick and pathogen life cycles (5-7).

While TBD represents an emerging public health crisis, timely and accurate surveillance is severely constrained by both the passive nature of reporting (8) and limitations of existing diagnostic methods (9). Indirect immunofluorescence assays (IFA) are widely employed laboratory tests for TBD which rely on detection of host antibodies to the infecting pathogen (10). Because antibodies are often not present at high levels during the first week of illness – the period when most patients initially seek care – a negative test cannot rule out disease. To confirm the diagnosis of SFGR or ehrlichiosis, a second or “convalescent” test is required two to ten weeks after the initial, “acute”, test. Unfortunately, few patients undergo both acute and convalescent testing with less than 3% of SFGR cases reported to the CDC being classified as confirmed (11). In the absence of paired acute and convalescent results, interpretation of a single titer is challenging, especially in areas where background rates of seropositivity are high (12).

In contrast, testing for Lyme disease employs a two-tier algorithm on a single serum sample, consisting of an enzyme immunoassay (EIA) or IFA, followed by a western immunoblot if the EIA is positive or equivocal (13). While this approach does not require multiple samples collected at different time points, real-world practice, especially in historically lower-incidence states, is fraught with ordering errors (i.e., western blot without a preceding EIA) and misinterpretation of results (14). Furthermore, incorporation of the much less specific IgM-based assay, intended to increase sensitivity early in the disease course, often increases false positivity rates (15, 16).

Diagnosis and prevention of TBDs depends greatly on an accurate understanding of local TBD epidemiology, much of which is derived from routine clinical practice, thus improving provider adherence to diagnostic testing guidelines remains a key priority (17). Without new testing methodologies or rigorous protocols to ensure high levels of testing completeness, estimates of disease incidence will remain susceptible to both under- and over-representation, the magnitude of which is ill-defined. Perhaps more importantly, misunderstanding of the predictive value of test results, especially a negative acute result, may contribute to delays in diagnosis and treatment, which is one of the main risk factors for severe disease and death (18-20).

To date, there has been a relative paucity of research examining the patterns of diagnostic testing for tick-borne disease, particularly for SFGR and ehrlichiosis. Moreover, previous studies have largely focused on data derived from routine surveillance, which does not include individuals who test negative. Including individuals who test negative provides important information about the underlying population assumed to be at risk and can yield complementary information regarding disease incidence and routine clinical practice. Therefore, the overarching objective of this study was to explore patterns in diagnostic testing for TBD in a large academic health system with the goals of (i) describing the demographic characteristics of patients tested for TBD, (ii) determining rates of adherence to testing guidelines and (iii) identifying potential gaps in provider ordering behaviors that may be improved with systems-based interventions (e.g., panels, reflex testing).

## Methods

### Study Site

North Carolina (NC) experiences some of the highest rates of SFGR and ehrlichiosis in the United States, often accounting for more than of 10% and 5% of cases reported to the CDC, respectively (11). In 2019, the incidence rate of SFGR was 6.6 cases per 100,000 residents with most cases being geographically clustered in the central and eastern parts of the state (21). Ehrlichiosis is less frequently reported, but is an under-recognized cause of tick-borne illness, despite evidence that infection rates may be similar to those of SFGR (18). Cases of Lyme disease are increasing in the state and are more concentrated in northwestern parts of the state in proximity to the Appalachian Mountains (22, 23).

We obtained diagnostic test results from the Carolina Data Warehouse for Health, a central repository containing clinical, research, and administrative data sourced from UNC Health. UNC Health is the largest academic health system in North Carolina, comprised of 12 hospitals and 350 outpatient clinics located across the state. In 2018, UNC Health reported approximately 3.5 million clinical visits including nearly 500,000 emergency department visits.

### Data Sources, Management, and Analysis

We requested the results of diagnostic testing performed by UNC’s McLendon Clinical Laboratories for SFGR, ehrlichiosis, and Lyme disease for the period from January 1^st^, 2017 to November 30^th^, 2020. Specifically, we abstracted results of IFA testing for immunoglobulin G (IgG) antibodies against *R. rickettsia* and *E. chaffeensis*, recognizing that cross-reactivity between species is well-known (24). Immunoglobulin M (IgM) results for *Rickettsia* or *Ehrlichia*, when ordered, were excluded from the analysis given that these tests are not included in the current testing guidelines. For Lyme disease, we abstracted the results of EIA, western blots and polymerase chain reaction (PCR). Western blot results were included in the analysis regardless of the preceding EIA result and even if the EIA was not ordered or not available. For each result, we documented the date of the test and age, sex, and race/ethnicity of the patient. Data were cleaned to remove duplicate entries. All analysis was performed using R statistical software (R Core Team, 2020). P-values for seasonal differences, defined as high season (March – October) and low season (November – February) based on historical tick activity [36], are based on the null assumption of equal testing throughout the year.

### Case Definitions

We established operational case classifications and laboratory criteria (**Table 1**) modeled after the CDC’s 2020 Surveillance Case Definitions used for SFGR and ehrlichiosis (25, 26). We excluded clinical criteria from these definitions due to lack of clinical information. Tests for SFGR and ehrlichiosis were considered paired if the acute and convalescent samples were performed on the same individual between 14 and 70 days of each other (25). Paired tests were subsequently defined to represent an incident case if either (i) the result of the acute test was ≤1:64 and the result of the convalescent test was ≥1:64 for ehrlichiosis or ≥1:128 for SFGR or (ii) the results of both acute and convalescent test were ≥1:64, and the convalescent result represented at least a four-fold increase in titer compared to the acute result. Paired tests were defined as a prevalent case if the convalescent titer was less than four-fold that of the acute and (i) both tests were ≥1:64 for ehrlichiosis or (ii) both tests were ≥1:64 with at least one test ≥1:128 for SFGR. In addition, we defined an individual as a probable case if at least one titer dilution was ≥1:128 or a suspected case if at least one test was positive at 1:64. A patient was considered a Lyme disease case if (i) their EIA test result was indeterminate or positive and the western blot satisfied criteria for either IgG or IgM positivity, or (ii) they lacked an antibody test performed and their western blot was positive for IgG, or (iii) they had a positive PCR result from cerebrospinal fluid (CSF).

**Table 1.** Operational usage of diagnostic terminology and case classifications for SFGR and ehrlichiosis test results used throughout the study

| Term | Operational definition |
| --- | --- |
| Paired test | Acute and convalescent IFA testing performed within 14-70 days |
| Incident case | Paired tests with four-fold increase in titer from acute to convalescent sample<br>OR<br>Paired tests with change from negative (<1:64) to positive (≥1:64 for ehrlichiosis, ≥1:128 for SFGR) from acute to convalescent sample; also referred to as seroconversion |
| Prevalent case | Paired positive tests with less than four-fold change in titer between acute and convalescent test, both tests ≥1:64 for ehrlichiosis, and at least one test ≥1:128 for SFGR |
| Suspected case | At least one positive test result at ≥1:64 |
| Probable case | At least one positive test result at ≥1:128 |
| Negative | No positive test result at ≥1:64 |

Abbreviations: IFA = indirect fluorescent antibody

### Ethical Approval

The study was approved by the institutional review board of the University of North Carolina at Chapel Hill (IRB 20-3502). As a limited data set under CFR 45, Part 164.514 (e), written informed consent or waiver of HIPAA authorization was not required.

## Results

During the study period, a total of 20,528 diagnostic tests for TBD were performed on 11,367 unique individuals, including 11,977 tests for Lyme disease from 10,208 individuals, 5,448 tests for SFGR from 4,520 individuals, and 3,103 tests for ehrlichiosis from 2,507 individuals (**Table 2**). More than one-third (n=4,144, 36.5%) of individuals were tested for more than one TBD with combinations Lyme, SFGR, and ehrlichiosis (n=1,724, 15.2%) and Lyme and SFGR (n=1,686, 14.8%), being most frequent. Testing for SFGR and ehrlichiosis was significantly higher during high season (March – October) with 88.0% (p<.001) and 86.5% (p<.001) of tests occurring during this period, respectively (**Figure 1**). In contrast, testing for Lyme was more equally distributed throughout the year, (p=.052). When stratified by the year testing was performed (**Table 3**), we observed a large decline in total tests performed in 2020, even when accounting for missing December 2020 data.

**Table 2:** Demographics for individuals tested and testing positive for tick-borne disease within the UNC Health system, 2017-2020.

|  | Overall | Lyme |  | SFGR |  | Ehrlichiosis |  |
| --- | --- | --- | --- | --- | --- | --- | --- |
|  |  | Tested | Positive | Tested | Incident | Tested | Incident |
| **Individuals, n** | 11367 | 10208 | 76 | 4520 | 47 | 2507 | 27 |
| **Sex** |  |  |  |  |  |  |  |
| Male, n (%) | 4734 (41.6) | 4150 (40.7) | 36 (47.4) | 2137 (47.3) | 18 (38.3) | 1206 (48.1) | 13 (48.1) |
| Female, n (%) | 6633 (58.4) | 6058 (59.3) | 40 (52.6) | 2383 (52.7) | 29 (61.7) | 1301 (51.9) | 14 (51.9) |
| **Age median (IQR)** | 53 (37-66) | 53 (38-66) | 46 (30-62) | 52 (36-66) | 58 (45-68) | 53 (38-67) | 61 (38-73) |
| **Race** |  |  |  |  |  |  |  |
| White, n (%) | 8850 (77.9) | 7993 (78.3) | 59 (77.6) | 3656 (80.9) | 40 (85.1) | 2027 (80.8) | 25 (92.6) |
| Black/African-American, n (%) | 1282 (11.2) | 1137 (11.1) | 8 (10.5) | 408 (9.0) | 3 (6.4) | 217 (8.7) | 0 (0.0) |
| Asian, n (%) | 116 (1.0) | 101 (9.9) | 0 (0.0) | 45 (1.0) | 1 (2.1) | 24 (1.0) | 0 (0.0) |
| American Indian/Alaska Native, n (%) | 56 (0.5) | 49 (0.5) | 0 (0.0) | 0 (0.0) | 0 (0.0) | 14 (0.6) | 0 (0.0) |
| Other, n (%) | 686 (6.0) | 591 (5.8) | 3 (3.9) | 277 (6.1) | 1 (2.1) | 161 (6.4) | 1 (3.7) |
| Unknown, n (%) | 377 (3.3) | 337 (3.3) | 6 (7.9) | 134 (3.0) | 2 (4.3) | 64 (2.6) | 1 (3.7) |
| **Ethnicity** |  |  |  |  |  |  |  |
| Hispanic/Latino, n (%) | 574 (5.0) | 494 (4.8) | 3 (3.9) | 242 (5.4) | 2 (4.3) | 140 (5.6) | 1 (3.7) |
| Not Hispanic/Latino, n (%) | 10288 (90.5) | 9272 (90.8) | 68 (89.5) | 4096 (90.6) | 44 (93.6) | 2273 (90.7) | 25 (92.6) |
| Unknown, n (%) | 505 (4.4) | 442 (4.3) | 5 (6.6) | 182 (4.0) | 1 (2.1) | 94 (3.7) | 1 (3.7) |

Abbreviations: IQR = interquartile range

**Table 3:** Annual trends in diagnostic testing and test positivity for tick-borne disease

|  | Lyme |  | SFGR |  |  | Ehrlichiosis |  |  |
| --- | --- | --- | --- | --- | --- | --- | --- | --- |
|  | Total | TPR (%) | Total | TPR (%) | Incident Cases | Total | TPR (%) | Incident Cases |
| **2017** | 3727 | 4.5 | 1621 | 32.9 | 5 | 724 | 16.7 | 3 |
| **2018** | 3576 | 3.0 | 1939 | 31.9 | 5 | 1590 | 36.1 | 10 |
| **2019** | 3415 | 4.5 | 1471 | 54.0 | 13 | 1100 | 47.6 | 7 |
| **2020** | 2691 | 4.1 | 1135 | 63.9 | 24 | 978 | 53.2 | 7 |

Abbreviations: TPR = test positivity rate

**Figure 1:**
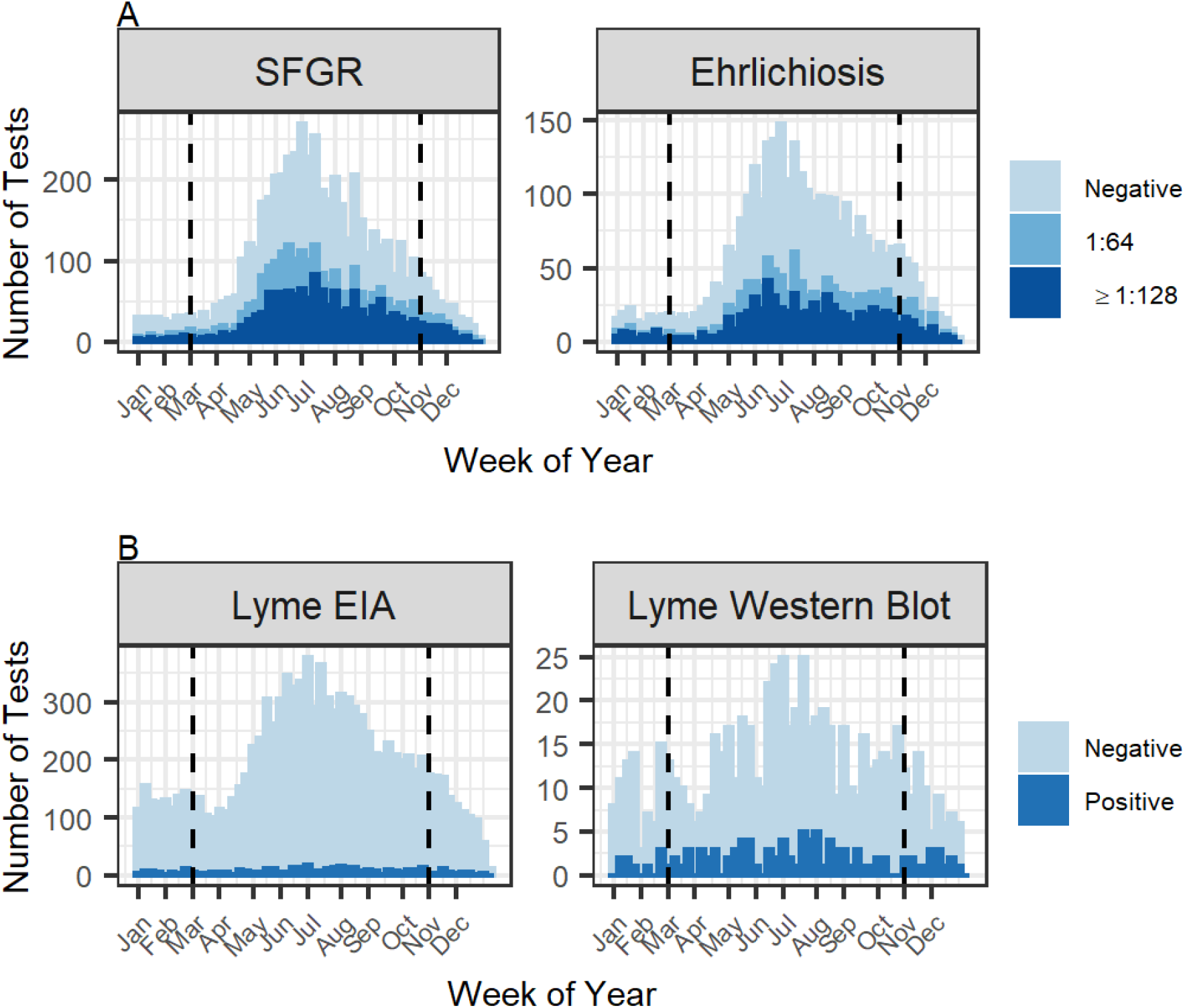
Seasonal trends of testing and positive tests for SFGR and Ehrlichiosis (1A) and Lyme Disease (1B) in NC from January 1^st^, 2017 to November 30^th^, 2020. Tests are aggregated by week of the year, with weekly number of tests given by the height of the bar and test result denoted by the color. Tick season (March – October) is designated by the vertical dashed line. For Lyme disease, equivocal EIAs are included with positive results.

The median age of individuals tested was 53 years (interquartile range [IQR] 37 - 66). Children under 18 represented 5.9% of those tested, much lower than the system-wide under 20 patient population in 2019 (27). Women represented 58.4% of individuals tested, similar to the demographic profile of patients system-wide, while 90.5% identified as non-Hispanic (83.1% system-wide) and 77.9% as white (60.8% system-wide). Racial and ethnic breakdowns did not vary substantially between those tested for each disease.

The overall test positivity rate (TPR) for SFGR, defined as the proportion of all tests which were positive at ≥1:128, was 27.1%, and for ehrlichiosis the TPR was 38.8%, reflecting positive results ≥1:64. The TPR for Lyme, defined as the proportion of positive EIA, PCR, and western blot results, was 4.2%. TPR increased substantially over the period of analysis for SFGR and for ehrlichiosis, which more than doubled from 16.7% to 53.2%. In contrast, TPR for Lyme remained relatively stable. Among individuals with paired tests, there were 25 (4.7%) incident cases of SFGR, and 27 (7.1%) incident cases of ehrlichiosis (**Table 4**). A total of 76 (0.7%) individuals satisfied the two-tier testing criteria for Lyme disease, among which 11 met both IgM and IgG criteria, 12 met only IgG criteria, and 53 met only IgM criteria (**Table 5**). Of the 224 Lyme PCR tests conducted on CSF, none were positive. TPR was similar between seasonal periods for Lyme but was higher during high season for SFGR (p=.01) and during the low season for ehrlichiosis (p<.01).

**Table 4:** Number of tests and paired testing completeness, SFGR and ehrlichiosis

|  | SFGR | Ehrlichiosis |
| --- | --- | --- |
| **Individuals** | 4520 | 2507 |
| *Individuals with paired tests, n (%)* | 536 (11.8) | 378 (15.1) |
| Incident case | 25 (4.7) | 27 (7.1) |
| Seroconversion | 19 (76.0) | 17 (63.0) |
| 4-fold increase | 6 (24.0) | 10 (37.0) |
| Prevalent case | 267 (49.8) | 239 (63.2) |
| Probable case | 626 (13.8) | 384 (15.3) |
| Suspected case | 288 (6.4) | 233 (9.3) |
| **Total tests** | 5448 | 3103 |
| Positive tests, n (%) | 2396 (44.0) | 1206 (38.8) |
| 1:128 threshold | 1475 (61.6) | 290 (24.0) |
| No paired test | 1517 (27.8) | 640 (53.1) |
| Negative tests, n (%) | 3029 (55.6) | 1897 (61.1) |
| Equivocal | 23 (0.4) | 0 |
| Paired tests (total) | 1128 (20.7) | 797 (25.7) |
| Number of tests without a convalescent pair, n (%) | 4320 (79.3) | 2306 (74.3) |

**OLD TABLE:**
|  | SFGR | Ehrlichiosis |
| --- | --- | --- |
| **Individuals** | 4520 | 2507 |
| *Individuals with paired tests, n (%)* | 536 (11.8) | 378 (15.1) |
| Incident case (1:64) | 47 (8.8) | 27 (7.1) |
| Seroconversion | 35 (74.4) | 17 (63.0) |
| 4-fold increase | 12 (25.5) | 10 (37.0) |
| Prevalent case (1:64) | 343 (64.0) | 239 (63.2) |
| Incident Case (1:128) | 25 (4.7) | 14 (3.7) |
| Seroconversion | 19 (76.0) | 11 (78.5) |
| 4-fold increase | 6 (24.0) | 3 (21.4) |
| Prevalent case (1:128) | 243 (45.3) | 157 (41.5) |
| Probable case | 626 (13.8) | 384 (15.3) |
| Suspected case | 288 (6.4) | 233 (9.3) |
| **Total tests** | 5448 | 3103 |
| Positive tests, n (%) | 2396 (44.0) | 1206 (38.8) |
| 1:128 threshold | 1475 (61.6) | 290 (24.0) |
| No paired test | 1517 (27.8) | 640 (53.1) |
| Negative tests, n (%) | 3029 (55.6) | 1897 (61.1) |
| Equivocal | 23 (0.4) | 0 |
| Paired tests (total) | 1128 (20.7) | 797 (25.7) |
| Number of tests without a convalescent pair, n (%) | 4320 (79.3) | 2306 (74.3) |

**Table 5:**
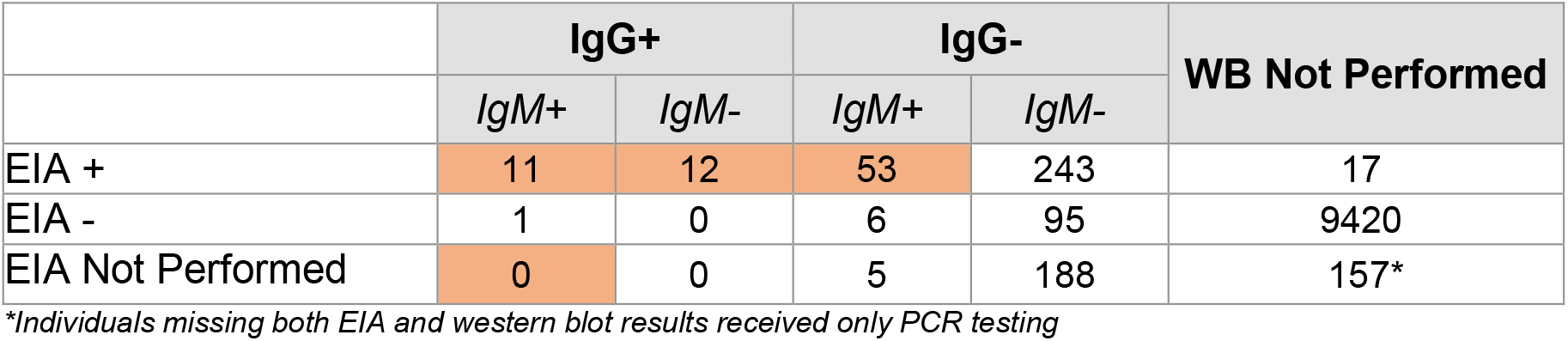
Serial testing results for individuals tested for Lyme disease, enzyme immunoassay (EIA) and western blot (WB)

Additionally, testing identified 287 prevalent, 626 probable, and 288 suspected cases of SFGR and 239 prevalent, 384 probable, and 233 suspected cases of ehrlichiosis. In total, 28.8% of individuals tested for SFGR and 35.2% of individuals tested for ehrlichiosis met the laboratory case definition of a suspected or probable case. Compared to all individuals tested, incident cases of SFGR and ehrlichiosis tended to be slightly older, while confirmed cases of Lyme tended to be younger and male (**Table 2**).

Of the 4,577 individuals that were tested for SFGR, only 536 (11.8%) had paired acute and convalescent testing performed. A modestly higher rate of paired testing was observed with ehrlichiosis (378 of 2507, 15.1%) (**Table 4**). Individuals with paired testing tended to be older (median age 57 vs 52 for SFGR; 56 vs 53 for ehrlichiosis) than those without paired testing. Additionally, individuals with an acute test result at ≥1:64 were more than twice as likely to have a convalescent test ordered. Adherence to standard testing protocol was higher for Lyme disease, with only 295 individuals (2.9%) undergoing a western blot without a preceding indeterminate or positive antibody test. Of these, 12 (4.1%) had positive IgM results and 1 (0.3%) had a positive IgG result (**Table 5**).

If individuals who only received an acute test experienced the same incidence rate as those with paired testing, we estimate that approximately 187 incident cases of SFGR and 151 incident cases of ehrlichiosis were not identified as a result of incomplete testing. Furthermore, because SFGR and ehrlichiosis have overlapping clinical disease spectra – and thus individuals presenting with typical symptoms (e.g., fever, headache, myalgia) and/or suspected tick exposure should be tested for both diseases – we estimate that there were 2,256 “missing” ehrlichiosis tests. Again, assuming similar incidence rates among individuals in whom SFGR was considered, but testing for ehrlichiosis was not performed, there would be an additional 158 “missing” incident cases, raising the estimate of total missed ehrlichiosis cases to 309.

## Discussion

Despite increasing awareness of TBD as a growing public health threat, our study demonstrates that critical gaps remain in the routine surveillance systems underlying our knowledge of the spatial and clinical epidemiology of these diseases. The low rate of testing completeness, with only one in ten individuals tested for SFGR having both acute and convalescent testing performed, severely limits the ability to distinguish incident infection from prior exposure. The paucity of confirmed diagnoses, both positive and negative, confounds prevention and control efforts, and introduces substantial misclassification bias into epidemiological and clinical research. Of particular concern, the apparent failure to consider ehrlichiosis as a potential cause of acute illness in nearly half of patients tested for SFGR raises serious doubts about our understanding of the burden of this disease. Improving the quality of routine surveillance data, through targeted education, systems-based interventions, and new testing modalities that do not require multiple visits, remains a key priority.

There are multiple potential explanations for the low level of testing completeness, including: (i) the resolution of clinical symptoms, either due to treatment or self-limited disease, that might reduce a patient’s motivation to return for a convalescent test, especially if incurring out-of-pocket costs (ii) the confirmation of alternative causes of illness over the interval between acute and convalescent testing, and (iii) a general lack of frontline provider knowledge regarding testing algorithms. One key to overcoming these issues is the development and more widespread adoption of acute stage diagnostics such as PCR. We note that the Food and Drug Administration recently approved a pan-*Rickettsia* real-time PCR assay and laboratory-developed assays are available for *Ehrlichia*, but these tests are often inaccessible to frontline providers and even when available remain underutilized for reasons that merit further investigation.

While rates of testing completeness were relatively low - although still higher than similar data at the state or national level - our results identified notable trends in both testing and infection. First, the overall cohort here was relatively older and more female than previous national surveillance studies (28, 29), although these characteristics are generally similar to the larger population seeking care at UNC Health. We did, however, observe a contrast in the demographics of individuals testing positive for each disease. Patients testing positive for Lyme disease were younger and more male, whereas those testing positive for SFGR and ehrlichiosis trended older and more female than those testing negative for these diseases. Whether these trends are due to differences in exposure risk, age-related clinical manifestations, or care-seeking behaviors is unclear. It is plausible that given the high levels of mild and even asymptomatic seroconversion previously seen in relatively young outdoor workers (24, 30), we may be observing differential care seeking patterns. In other words, infection with less pathogenic bacteria such as *R. parkerii* or *R. amblyommatis* may result in mild or asymptomatic disease in younger individuals, but more apparent symptoms in older individuals that subsequently prompts care-seeking and testing.

Another crucial finding is the high rate of testing for Lyme disease in contrast to the relative neglect of testing for ehrlichiosis, even among those tested for SFGR. The low rate of testing completeness and lack of consideration of ehrlichiosis likely resulted in a large number of unidentified cases, which would greatly impact surveillance data. While North Carolina reported a total of 1,704 cases of SFGR and 372 cases of ehrlichiosis statewide over the study period, our estimates suggest there might be an additional 187 SFGR and 309 ehrlichiosis cases from the UNC Health system alone. Our estimates, however, assume that individuals without paired testing experienced the same incidence rate as those with paired testing. These groups, though, differ significantly in ways that may affect the likelihood of a positive convalescent test and ultimately may overestimate the number of missed cases. In contrast, the high rate of Lyme testing may artificially inflate incidence rates in the area due to false positive results, especially with the IgM assay, which represented a majority (69.7%) of positive western blot results.

Our study has several strengths including the large sample size abstracted from both outpatient and inpatient settings distributed over a relatively large geographic area. Furthermore, unlike previous analyses of notifiable disease reports, our study includes information about individuals who tested negative; information not normally available in these reports. Lastly, with high rates of SFGR and ehrlichiosis, and expanding Lyme in the western regions, North Carolina is a compelling setting to monitor trends in patterns of TBD testing.

The current study also has important limitations. Foremost among these is the lack of clinical data such as the nature, onset and duration of symptoms that would have allowed us to more fully apply case definitions. Particularly, we are unable to identify cases in which initial testing was done within the first seven days of illness onset, important because antibodies to many TBDs may not yet be detectable. Second, the absence of longitudinal data limits our ability to assess treatment decisions and, most importantly, clinical outcomes. Lastly, our study population may not be wholly representative of all patients seeking care for TBD in the area. Some patients may have been empirically treated without formal diagnostic testing based on the presence of typical symptoms and exposure history alone. If individuals with “textbook” disease were more likely to have been empirically treated and less likely to be tested, this may have biased our sample.

## Conclusions

Our study showed that a majority of patients suspected of having TBD did not have testing performed in accordance with established guidelines, which substantially limits our understanding of TBD epidemiology. Furthermore, there appears to be a large discrepancy between the local burden of disease and the testing that is performed. These findings underscore the need for better diagnostics and active surveillance programs, particularly for SFGR and ehrlichiosis, to more accurately identify the spatial distribution and risk factors for infection.

## Data Availability

Deidentified individual data that supports the results will be shared beginning 9 to 36 months following publication provided the investigator who proposes to use the data has approval from an Institutional Review Board (IRB), Independent Ethics Committee (IEC), or Research Ethics Board (REB), as applicable, and executes a data use/sharing agreement with UNC.

## DECLARATIONS

### Conflicts of Interests

All authors have completed the ICMJE uniform disclosure form and declare: no financial relationships with any organizations that might have an interest in the submitted work in the previous three years except that noted in the funding section; no other relationships or activities that could appear to have influenced the submitted work.

### Previous Publication

The authors confirm that the submitted work has not been previously presented or published in any format and is not under consideration at any other journal.

### Funding

Funding for the study was provided by a Creativity Hub Award to RMB from the UNC Office of the Vice Chancellor for Research. RMB is also supported by a Caregivers at Carolina Award made by the Doris Duke Charitable Foundation (Award 2015213). Research was supported by the North Carolina Translational and Clinical Sciences (NC TraCS) Institute, which is supported by the National Center for Advancing Translational Sciences (NCATS), National Institutes of Health, through Grant Award Number UL1TR002489.

### Author Contributions

Study conception and design: ABM, BDH. Funding: RMB. Study implementation: BDH, ABM. Data analysis: BDH, ABM, RMB. First draft of manuscript: ABM, BDH, HA, AA, VA, OM. Revisions: All.

## Acknowledgements

We wish to thank the Regulatory team at the UNC Institute of Global Health and Infectious Diseases as well as the Data Management team at the Carolina Data Warehouse for their efforts.

## Notes

### Competing Interest Statement

The authors have declared no competing interest.

