## Supplemental Table for "“Half the truth is often a great lie:” testing for tick-borne diseases and implications for surveillance"

### SUPPLEMENTAL TABLES

**Table S1:** Total number of individuals tested for Lyme disease and testing results

| | Antibody | IgG | IgM | PCR | Overall |
|---|---|---|---|---|---|
| Individuals | 9858 | 614 | 614 | 222 | 10208 |
| Total Tests | 10460 | 666 | 666 | 224 | 12016 |
| Positive Tests | 197 (2.0) | 26 (4.2) | 88 (14.3) | 0 (0.0) | 311 (3.1) |
| Negative Tests | 10240 | 639 | 557 | 224 | 11680 |
| Equivocal | 23 | 1 | 1 | 0 | 25 |
| Confirmed Cases | --- | --- | --- | --- | 76 (0.8) |
